# Clinical impact of the *Predict* Prostate risk communication tool in men newly diagnosed with non-metastatic prostate cancer: a multi-centre randomised controlled trial

**DOI:** 10.1101/2021.01.24.21249948

**Authors:** David Thurtle, Val Jenkins, Alex Freeman, Mike Pearson, Gabriel Recchia, Priya Tamer, Kelly Leonard, Paul Pharoah, Jonathan Aning, Sanjeev Madaan, Chee Goh, Serena Hilman, Stuart McCracken, Petre Cristian Ilie, Henry Lazarowicz, Vincent Gnanapragasam

## Abstract

**Introduction:** Predict *Prostate* is a freely-available online personalised risk communication tool for men newly diagnosed with non-metastatic prostate cancer. Its accuracy has been assessed in multiple validation studies but the clinical impact of the tool on patient decision-making had not previously been evaluated.

**Methods:** A multi-centre randomised controlled trial was performed across 8 UK centres, wherein newly diagnosed men considering either active surveillance or radical treatment, were randomised to either standard of care (SOC) information or SOC and presentation of Predict *Prostate*. Validated questionnaires were completed assessing impact of the tool on decisional conflict, uncertainty, anxiety and understanding of survival.

**Results:** 156 patients were included; mean age 67 years (range 44-80) and PSA of 6.9ng/ml (range 0.5-59.8). 81 were randomised to the Predict *Prostate* arm, and 75 to SOC information only. Mean decisional conflict scores were 26% lower in the Predict *Prostate* group (mean = 15.9) than in the SOC group (mean = 21.5) (p=0.01). Scores on the ‘effective decision’, ‘uncertainty’ and ‘value clarity’ subscales all indicated that the Predict *Prostate* group felt more informed and clear about their decision (all p<0.05). There was no significant difference in anxiety between the two groups.

Patient perceptions of 15-year prostate cancer specific mortality (PCSM) and overall survival benefit from radical treatment were considerably lower among men in the Predict *Prostate* group (p<0.0001). 58% of men reported the Predict *Prostate* estimates for PCSM were lower than expected, and 35% reported being less likely to select radical treatment. Over 90% of patients in the Predict *Prostate* group found it useful and 94% would recommend it to others.

**Conclusion:** Predict *Prostate* reduces decisional conflict and uncertainty in non-metastatic prostate cancer and shifts patient perceptions around prognosis to be more realistic. This is the first randomised study of such a tool in this context; it demonstrates Predict *Prostate* can directly inform the complex decision-making process in prostate cancer.

## BACKGROUND

The majority of prostate cancer (PCa) presents with non-metastatic localised or locally advanced disease, equating to over 36,000 men in the UK (1). Treatment decisions are notoriously complex, particularly for men with earlier stage disease with the risk of progression and psychological impact of a cancer diagnosis balanced against potential morbidity associated with radical treatment. Unsurprisingly, decisional anxiety and regret are well-recognised issues for many newly diagnosed men (2). There is also significant regional variation in the proportion of men undergoing radical treatment, particularly for favourable intermediate-risk disease (3). Prognostic stratification is therefore vital in guiding management and treatment decision-making.

Predict *Prostate* (prostate.predict.nhs.uk) is a freely-available online personalised risk communication tool based on an internationally-validated prognostic model for men with newly diagnosed, non-metastatic PCa (4-6). It has been endorsed by the UK National Institute for Health and Care Excellence (NICE) and is the only such tool CE-marked for use in clinical decision-making in PCa (4). Using details on patient characteristics and clinic-pathological features, the tool produces personalised PCa-specific and overall survival estimates displayed in a range of visual outputs to contextualise the potential benefits of radical treatment. Potential harms of the different treatment options are also presented using data from published series. The modelling and tool was designed to meet all American Joint Committee on Cancer (AJCC) criteria for model adoption, and has been demonstrated to outperform existing risk-models, with concordance indices in excess of 0.8 in a number of international datasets (6, 7). The interface was based on a design with extensive input from patients and quantitative testing in order to improve risk comprehension and support decision-making without being ‘persuasive’ to patients in a particular way (8). Previous work has demonstrated the tool impacts upon clinician perceptions and treatment recommendations (9). However, to date the clinical utility of such a risk communication tool embedded within an interactive web interface has not been tested with newly diagnosed PCa patients.

Impact analyses are an important element of prognostic model development and should include comparisons to a control group who receive standard care (10). Impact studies can also be useful to study issues that may affect acceptability and uptake of a model in regular care as well as usability (10). In this impact study, we assessed whether or not using Predict *Prostate* materially affected patient decision-making within a multi-centre randomised controlled trial design. Our primary objectives were to assess the impact of the Predict Prostate risk communication tool on decisional conflict, uncertainty and anxiety amongst men newly diagnosed with non-metastatic PCa. Secondary objectives were to assess how Predict *Prostate* survival estimates compared to and impacted upon patients’ perceptions of their survival and survival benefits of treatment, retrieve feedback on the tool and assess whether certain men may benefit more from using the tool.

## STUDY DESIGN AND PARTICIPANTS

### Trial design

The study was an ethically approved (REC 18/EE/0254) UK multi-centre prospective randomised study. The full protocol is available online (ISRCTN 28468474)(11). In summary, men aged 35-80 with newly diagnosed non-metastatic PCa, in whom either surveillance or upfront radical treatment were both deemed potentially appropriate, were invited to participate (Supp Table S1). Study participation was integrated into the patient’s clinical pathway, without delays in standard management pathways. Participants, already aware of their PCa diagnosis were invited to a study appointment. All participants were informed and counselled according to the SOC in their centre’s normal practice, which we did not standardise across centres. During the study appointment men were randomised to either the control (standard of care (SOC)) arm or the intervention (Predict *Prostate*) arm of the trial. Randomisation was achieved by block random allocation within each site (with random block sizes between 4-6)(*12*). Recruitment took place between November 2018 and March 2020. Men in the SOC arm were directly allocated to complete the questionnaire. Those in the intervention arm were exposed to a structured presentation of the Predict *Prostate* tool by a trained researcher, then completed the questionnaire. The researchers presented the tool by directing the participant through the website without offering any additional clinical advice or input, using prescribed terminology (Additional File 1).

### Assessment measures

The questionnaires are available in Additional File 2. Validated scoring systems were used, namely the Decisional Conflict Scale (DCS), Decision Making Preference Questionnaire (DMPQ) and State-Trait Anxiety Inventory (STAI) (13-15). The importance of various factors in patient decision-making such as survival, bowel function and sexual dysfunction were assessed using methodology published previously (16). Patient demographic and tumour details, perceptions around prognosis, and final treatment decision were also collected. Patients in the intervention arm completed additional questions on the model and were given the option to provide written feedback.

### Sample size estimate and statistical analysis

A priori information of impact on DCS and an accepted size of impact deemed clinically relevant was not available. Mirroring outcomes from a study using changes in actual treatment choices, we estimated that a reduction of DCS of 20% would be a clinically meaningful outcome (17). The minimum required sample size (SD 20, α 0.05, β 0.80) was therefore 32. To capture potential lower impact results, we expanded our target sample size to 150, which would detect DCS reductions of 10%. The impact of the intervention was tested within an intention-to-treat analysis. DCS, DCS subscales, and STAI scores were compared between groups using independent samples t-test, as pre-specified in the protocol. Data analyses were performed in Stata™ 14 (StataCorp, Texas, US). Participants’ feedback and comments were collated and summarised.

## RESULTS

### Patient characteristics

156 patients were included from 8 UK centres, representing a mixture of larger academic centres and smaller district general hospitals (Table 1). 81 were randomised into the Predict *Prostate* arm of the study and 75 to the SOC arm. Distribution of characteristics across the two arms were similar; median age was 67 years and PSA 6.9ng/L (Table 2). Other patient and tumour characteristics were predominantly consistent with low or intermediate-risk disease, according to NICE or EAU criteria (Table 2)(18). More than 99% of patients were diagnosed following a pre-biopsy MRI and the majority had both targeted and systematic prostate biopsies. Transperineal biopsies were performed in 56.3% of cases (Supp Table S2). Additional educational and social characteristics of the patients are recorded in Supp Table S3, with no significant differences between the groups. Only 11/156 (7%) patients had exposure to the Predict *Prostate* tool prior to partaking in the study, (intervention group n=6; control group n=5).

**Table 1:** Recruiting centres and number of men completing the trial at each site.

| **Centre** | **Completed study** |
| --- | --- |
| City Hospitals Sunderland, Sunderland | 6 |
| Cambridge University Hospitals, Cambridge | 38 |
| Dartford and Gravesham, Dartford | 39 |
| North Bristol, Bristol | 40 |
| Queen Elizabeth Hospital, Kings Lynn | 4 |
| Surrey and Sussex Healthcare, Redhill | 18 |
| Weston General Hospital, Weston-Super-Mare | 7 |
| Royal Liverpool and Broadgreen, Liverpool | 4 |
| **Total** | **156** |

**Table 2:** Patient and tumour characteristics within the intervention and SOC arm, and overall. Comparison is made between study arms. SOC = standard of care PSA = Prostate-Specific Antigen cT stage = clinical tumour stage

| Predict *Prostate* arm |  |  |  | SOC arm |  |  | Overall |  |  |  |  |
| --- | --- | --- | --- | --- | --- | --- | --- | --- | --- | --- | --- |
| (n=81) |  |  |  | (n= 75) |  |  | (n=156) |  |  |  |  |
| Variable | Med | Mean | SD | Med | Mean | SD | Med | Mean | SD | Min | Max |
| Age | 67 | 65.5 | 7.7 | 67 | 66.1 | 6.6 | 67 | 66.0 | 7.13 | 44 | 80 |
| PSA | 6.7 | 7.2 | 3.1 | 7.3 | 8.6 | 7.2 | 6.9 | 7.82 | 5.45 | 0.52 | 59.8 |
| Biopsy Cores taken | 16 | 17.4 | 9.5 | 14.5 | 18.4 | 8.8 | 16 | 17.9 | 9.15 | 1 | 52 |
| Biopsy cores positive | 4 | 5.1 | 5.0 | 4 | 5.3 | 5.2 | 4 | 5.2 | 5.1 | 1 | 27 |
|  |  | n | % |  | n | % |  | n | % |  |  |
| Grade Group | 1 | 34 | 42.0 |  | 27 | 36.0 |  | 61 | 39.1 |  |  |
|  | 2 | 38 | 46.9 |  | 39 | 52.0 |  | 77 | 49.4 |  |  |
|  | 3 | 7 | 8.6 |  | 8 | 10.7 |  | 15 | 9.6 |  |  |
|  | 4 | 2 | 2.5 |  | 1 | 1.3 |  | 3 | 1.9 |  |  |
| cT stage | cT1 | 24 | 30.3 |  | 19 | 26.4 |  | 43 | 28.5 |  |  |
|  | cT2 | 50 | 63.3 |  | 48 | 66.7 |  | 98 | 64.9 |  |  |
|  | cT3 | 5 | 6.3 |  | 5 | 6.9 |  | 10 | 6.6 |  |  |
| Risk Category | Low | 26 | 32.1 |  | 26 | 34.7 |  | 52 | 33.3 |  |  |
|  | Int | 49 | 60.5 |  | 44 | 58.9 |  | 93 | 59.6 |  |  |
|  | High | 6 | 7.4 |  | 5 | 6.7 |  | 11 | 7.1 |  |  |

### Decision-making preferences

Most patients wanted decision-making to be a collaborative process, with 55% preferring to ‘make treatment decisions with their doctor’ and 38% preferring to ‘make decisions after hearing their doctors’ opinion’ (Supp. Table S4). Dying from PCa was reported to be either an ‘important’ or ‘very important’ factor when decision-making by 84% men. Incontinence (77.4%) and bowel problems (78.7%) were also considered important; fewer felt that sexual dysfunction (51.3%) or the burden of treatment itself (57.4%) were important (Table 3).

**Table 3:** Reported importance of factors in participants’ decision-making around treatment. Scored on a 5-point scale using the terms at the heading of each column. The final column combines ‘important’ and ‘very important’.

|  | *Not*<br>*importa*<br>*nt* | *Slightly*<br>*importa*<br>*nt* | *Mode-*<br>*rately*<br>*importa*<br>*nt* | *Important* | *Very*<br>*importan*<br>*t* | **Important**<br>**OR**<br>**Very**<br>**Important** |
| --- | --- | --- | --- | --- | --- | --- |
|  | n | n | n | n | n | % |
| Chance of dying from prostate cancer | 1 | 10 | 13 | 32 | 98 | **84.4** |
| Risk of bowel problems | 2 | 8 | 23 | 72 | 50 | **78.7** |
| Risk of urinary problems | 2 | 7 | 26 | 79 | 41 | **77.4** |
| Thought of living with untreated cancer | 8 | 22 | 26 | 35 | 64 | **63.9** |
| Burden of treatment itself | 11 | 21 | 34 | 62 | 27 | **57.4** |
| Risk of sexual problems | 25 | 12 | 38 | 46 | 33 | **51.3** |

### Impact of Predict Prostate on survival perceptions

Patient perceptions of 15-year PCSM, non-PCa mortality (NPCM) with conservative management, and survival benefit from radical treatment were all considerably lower among men who had seen the Predict tool (Table 4). This was most striking for PCSM where patient-perceived 15-year risk of mortality was 41.9% among men in the control group, and 21.7% in the PREDICT group (p<0.0001). This was despite there being no differences between the two groups in terms of prognosis, as demonstrated by mean Predict **Prostate** estimates (Table 4). Although the patient perceived PCSM and survival benefit from treatment were lower among those in the Predict group, they continued to be much higher than Predict *Prostate* estimates, whereas patients’ perceptions of NPCM were similar to Predict *Prostate* estimates.

**Table 4:** Comparison of perceived 15-year Prostate cancer (PCa) mortality, non-PCa mortality, and survival benefit from radical treatment for men in the standard of care (SOC) group and those who saw the Predict *Prostate* tool. Mean Predict Prostate estimates are also recorded and compared between each randomisation group. P values relate to the comparison using independent group t-test.

|  | **SOC**<br>% Mean (95%CI) | **Predict**<br>% Mean (95%CI) | p |
| --- | --- | --- | --- |
| **Chance of dying from PCa over 15 years** |  |  |  |
| **Patient perception** | 41.9 (35.6-48.3) | 21.7 (16.9-26.4) | <0.0001 |
| **Predict estimates** | 7.96 (6.9-9.0) | 7.15 (6.5-7.8) | 0.10 |
| **Chance of dying from other (non-PCa) causes over 15 years** |  |  |  |
| **Patient perception** | 49.8 (44.4-55.1) | 34.3 (28.6-40.0) | 0.0001 |
| **Predict estimates** | 34.2 (29.6-38.8) | 35.1 (30.2-39.8) | 0.60 |
| **Extra men alive following radical treatment (vs CM) over 15 years** |  |  |  |
| **Patient perception** | 51.3 (44.1-58.6) | 34.8 (27.3-42.2) | 0.0009 |
| **Predict estimates** | 3.8 (3.3-4.2) | 3.5 (3.1-3.8) | 0.12 |

Men in the Predict *Prostate* arm also reported that the mortality estimates differed from their expectations, especially for PCSM where 58% of men reported the estimates were lower than they expected. Only 6% reported the Predict estimates for PCSM were higher than they expected. Most men (73%) felt that the NPCM estimates were similar to their expectations (Figure 1).

**Figure 1:**
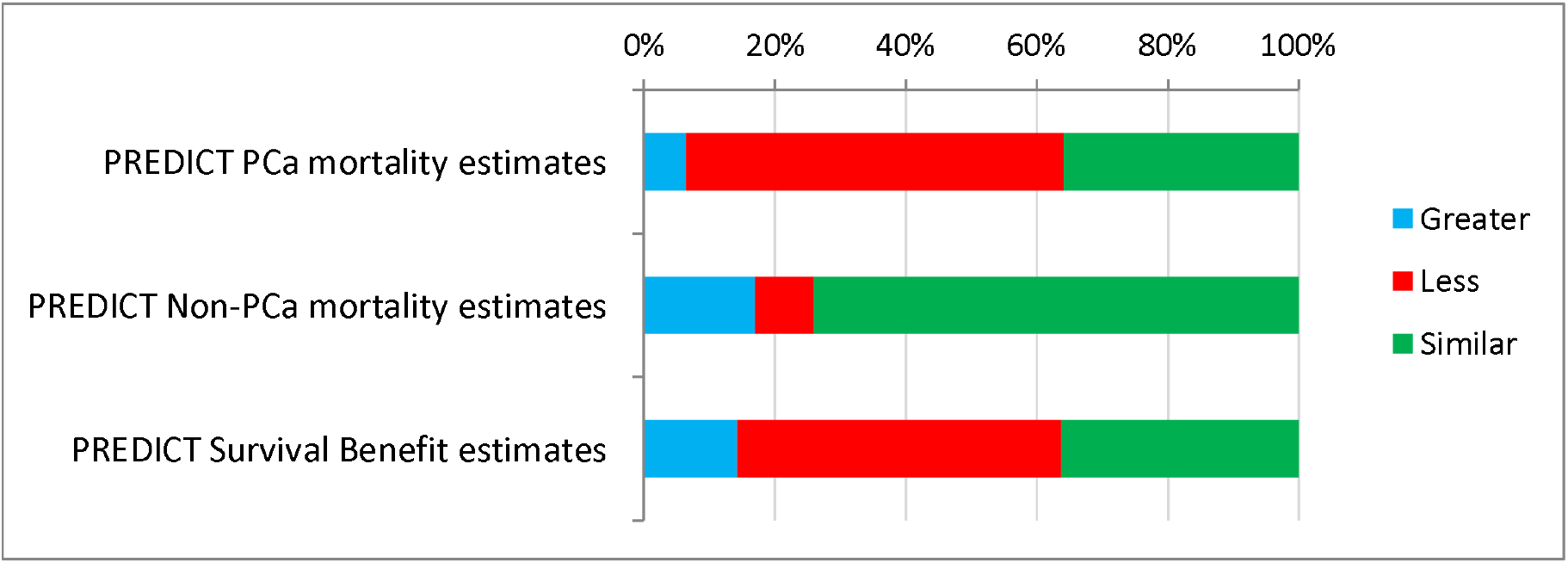
Participant-reported comparison of PREDICT estimates to their perceptions. Assessed only among the group shown the PREDICT model (n=78)

### Impact of Predict Prostate on decision-making confidence

Mean composite decisional conflict scores were 21.5 and 15.9 for the SOC and Predict groups respectively, representing 26% lower overall decisional conflict among those who saw the Predict *Prostate* tool (p=0.01) (Figure 2). This improvement was observed across nearly all of the DCS subscales including the ‘uncertainty’, ‘effective decision’ and ‘values clarity’ subscales (Table 5). There were no significant differences between either group in terms of trait (p=0.275) or state (p=0.124) of anxiety (Supp Table S5).

**Table 5:** Decisional conflict scale (DCS) and pre-defined subscale results for the control and Predict study arms. *p* values relate to the comparison using independent group t-test.

| **Measure** | **SOC (n=75)**<br>**Mean (95% CI)** | **Predict (n=81)**<br>**Mean (95% CI)** | **P value** |
| --- | --- | --- | --- |
| **DCS Informed Subscore** | 16.22(12.8-19.6) | 13.23(10.4-16.0) | 0.089 |
| **DCS Values Clarity Subscore** | 21.06(17.3-24.8) | 13.92(10.6-17.3) | 0.0027 |
| **DCS Support Subscore** | 15.88(11.7-20.0) | 10.44(7.7-13.2) | 0.0147 |
| **DCS Uncertainty subscore** | 32.42(27.0-37.8) | 24.18(19.0-29.3) | 0.0149 |
| **DCS Effective Decision Subscore** | 23.27(18.7-27.8) | 17.6(13.4-21.7) | 0.033 |
| **Decisional Conflict Scale Overall** | **21.54 (17.9-25.2)** | **15.91 (12.7-19.1)** | **0.01** |

**Figure 2:**
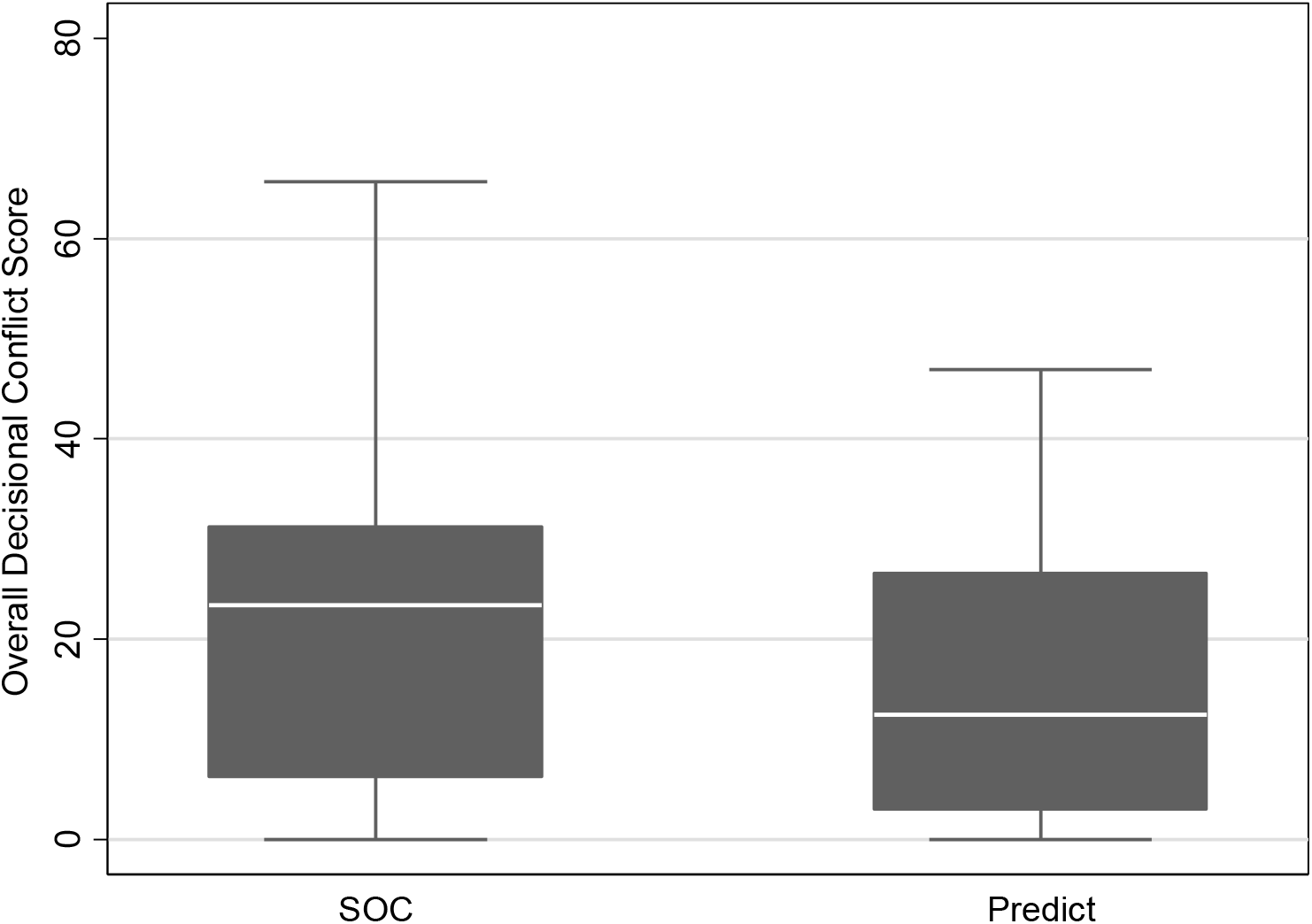
Comparison of mean overall decisional conflict scale results between the standard of care (SOC) group and Predict *Prostate* group (Predict). p=0.01.

When asked whether seeing the Predict model would make them more or less likely to choose radical treatment, 1 in 3 men (35%) reported it would make them less likely to choose radical treatment. 38% felt it would not change their decision, and only 15% felt Predict would make them more likely to choose radical treatment (Table 6). Overall, there were no differences in terms of reported treatment preferences (Supp Table S7) or the final treatment received between the two groups (Supp Table S9). However, Predict appears to have helped confirm patients’ treatment preferences, with over 80% of those who felt Predict made them less likely to want radical treatment opting for AS, and over 90% of those who felt the opposite opting for radical treatment (Supp Table S6).

### Patient feedback on Predict Prostate

90% of patients said they found Predict Prostate useful and 94% would recommend it to other men in their position (Supp Figures S1 + S2, Table S9). Free text responses and written feedback were overwhelmingly positive. Individual, anonymised, comments are recorded in Supp Table S10. Repeated themes were that the tool was ‘useful’, ‘informative’ and ‘helpful’.

## DISCUSSION

This study suggests that the Predict *Prostate* risk communication tool reduces decisional conflict when used in a clinical decision-making situation. The tool shifts patient perceptions around survival and treatment-benefit to be more realistic; and is popular with PCa patients. This randomised trial provides level 1 evidence with regards to the efficacy of the model in aiding treatment decisions and demonstrates it can be integrated into contemporary routine clinical practice without increasing patient anxiety.

Most existing literature has explored formal ‘decision aids’ rather than risk-communication tools such as Predict *Prostate*. Indeed we are unaware of any previous studies investigating using such tools and impact on patients. A well-conducted systematic review of decision aids for localised PCa, published in 2015, identified only 14 RCTs, none of which related to individualised or prognostic tools. The review concluded that ‘scant evidence at high risk of bias’ existed with regards to their benefit (19). The lack of RCTs of personalised models informing the decision between surveillance and radical treatment is presumably a result of an absence of these underlying nomograms until very recently.

Moons *et al* stated that the assumption that accurate outcome estimates lead to improved patient decision-making requires testing, with comparisons made to standard care (10). Although impact studies are an important part of model-development, few are performed on prognostic models, and fewer still have explored decision-making in PCa (20, 21). This study sought to address these inadequacies in the existing literature.

Our study confirms previous findings about PCa patients’ decision making preferences and the importance they place on various factors (16). Survival was reported to be the most important factor, perhaps justifying a prognosis-focussed tool such as Predict *Prostate* - built primarily around long-term survival. Sexual impact and treatment convenience were deemed less important (16). Our study shows that an individualised prognostic model can reduce decisional conflict by about 25%, and leads to similar improvements in terms of making an effective decision and clarifying values. It is reasonable to postulate that this will have positive sequelae in terms of engagement in shared decision-making and adherence to treatment regimes. Indeed, work with decision aids has shown this effect, with a large Cochrane review concluding that patients exposed to decision aids generally felt more knowledgeable, better informed and played a more active role in shared decision-making (22). As a result of feeling better-informed and engaged, more careful decisions may be made, which according to the Decision Justification Theory may reduce decisional regret – a common phenomenon in PCa patients (2, 23). Of note, decisional conflict in our study was higher than reported in a similar American cohort where mean DCS was 9.9 and 10.9 at 1 month following diagnosis, compared to 21.5 and 15.9 in our control and intervention groups respectively (24). Explanations for this difference may include patient characteristics, structural differences between the two nation’s healthcare systems, and possibly the higher number of consultations reported within the first month in that study (24).

Predict *Prostate* was also seen to reduce decisional uncertainty and there was no difference in anxiety between the groups, with a trend towards lower ‘state’ of anxiety among men in the Predict Prostate group. This finding should allay the concerns of some healthcare professionals that using models like these might increase uncertainty or anxiety among patients (25, 26).

We demonstrate that Predict *Prostate* also shifted perceptions around mortality between the two randomised groups. Individual patient-reported estimates of mortality among those in the control group were incredibly high, with a mean expectation that 42/100 of them would die from PCa within 15 years, and 51/100 extra men would survive with radical treatment. This is incongruous with RCT data from similar cohorts, such as the Pivot study where only 11.4% of men assigned to observational treatment died from prostate cancer over a median of 12.7 years, or the SPCG-4 trial where only 31.3% died from PCa after 23 years median follow-up (27, 28). Patients exposed to the Predict **Prostate** tool were still likely to have unrealistically high expectations of treatment benefit and disease lethality but perceptions of prognosis shifted significantly downwards towards more realistic values (Table 4). We previously demonstrated very similar findings among healthcare professionals in the PCa field, who consistently overestimated PCSM and survival benefits from treatment before using the tool, and in whom the likelihood of recommending radical treatment was often lower following exposure to Predict *Prostate* estimates (9). Again, this finding is in-keeping with conclusions from the Cochrane review which concluded decision-aids ‘probably’ led to more accurate risk perceptions amongst clinicians (22). With regards to survival estimates, we recognise Predict *Prostate* itself is not infallible, however, the tool has been externally validated in multiple cohorts and its calibration has been demonstrated to be accurate and superior to other available tools (5, 6).

The model appears to have face validity for patients, especially for NPCM where most reported the model estimates were similar to what they expected. A larger proportion of men in the Predict arm reported being less likely to choose radical treatment - presumably, due to the shift in perceptions on disease lethality. Other results also suggested the tool may help to solidify treatment preferences. The majority of those who felt the Predict tool made them more likely to choose radical treatment did so, and vice versa. Of note, we did not find any overall difference in the final treatment decision made between the groups but this study was not designed, and was underpowered, to answer this question.

Within the intervention group, no concerns were raised over usability of the tool, despite using a very diverse patient cohort by way of social characteristics. Open box feedback was overwhelmingly positive. Among those patients randomised to the Predict **Prostate** arm of the study, the model received positive feedback with over 90% reporting they found it to be useful, and that they would recommend it to others. It was not possible to assess whether particular patient sub-groups gained more from the model, due to the universally positive feedback.

Particular strengths of this study are the integration of this model within a contemporary PCa pathway, utilising up-to-date MRI and diagnostics. The model was tested within a randomised sample including a broad range of patients from multiple centres and disparate backgrounds representative of UK practice. We do also recognise the potential limitations of the study, including that patients and investigators could not be blinded to their randomisation group. Although we did not set out, nor power the study, to detect actual differences in treatment choices, we did observe there were no overall differences in treatment preference or final decisions between the groups. In this trial Predict Prostate was presented to patients in a delayed trial specific appointment, separate to their being informed about the diagnosis. We anticipate that the model would ordinarily be presented alongside counselling about the new diagnosis. This delay may have meant that men had already researched and decided upon the treatment options prior to seeing Predict Prostate. There was also a clear risk of contamination with regards this outcome as patients in the SOC arm may have subsequently accessed the online tool. These factors may therefore partly explain why we did not see actual differences in final treatment decisions between the groups, despite a third of men in the intervention arm saying they were less likely to have radical treatment after seeing the tool. Assessment outcomes of long-term survival or decisional regret, were also not measured but would be a useful comparison assessment in future studies. Whether the findings can be generalised beyond the UK healthcare setting is unclear, although the tool is accessed globally. Inclusion was also limited to patients able to read and write in English which may bias the results, and the tool remains untested in patients with reduced literacy. Future work could focus upon the optimal timing for delivery of the tool, its long term impact upon treatment practices, and its potential value among men within other healthcare settings.

## Conclusion

The Predict *Prostate* risk communication tool reduces decisional conflict and shift perceptions around prognosis without increasing anxiety and whilst being popular with most PCa patients. This work, alongside the pre-existing rigorous validation of the model should encourage health care professionals and patients alike to make use of the model to help inform what can be a complex decision-making process.

## Supporting information

Additional File 1 - Study appointment protocol

Additional File 2 - Patient Questionnaire

Additional File 3 - Study Protocol

## Data Availability

Formal requests for access to data should be made to the authors.

