## Supplementaries for "Clinical impact of the *Predict* Prostate risk communication tool in men newly diagnosed with non-metastatic prostate cancer: a multi-centre randomised controlled trial"

**Patient Study Supplementaries**

**Table S1 – Study eligibility criteria**

**Inclusion criteria**

- Men newly diagnosed with primary non-metastatic PCa.
- Men for whom either active surveillance or radical treatment (prostatectomy +/- radiotherapy) are felt to be appropriate by the diagnosing clinician.
- Age 35-80 years
- Able to understand and provide informed consent

**Exclusion criteria**

- Known to have a condition, which affects their ability to see, read or understand the decision aid
- Any other condition, which in the opinion of the investigator makes the subject unsuitable for study participation.
- Unable to comprehend English. (PREDICT Prostate is only available in English currently)

|  | **Predict *Prostate* arm** | | | **SOC arm** | | **Overall** | | |
| --- | --- | --- | --- | --- | --- | --- | --- | --- |
|  |  | n | % | n | % |  | n | % |
| **Biopsy route** | TP | 44 | 56.4 | 41 | 56.2 |  | 85 | 56.3 |
|  | TR | 34 | 43.6 | 32 | 43.8 |  | 66 | 43.7 |
| **Biopsy approach** | T&S | 36 | 45.0 | 43 | 58.1 |  | 79 | 51.3 |
|  | S only | 35 | 43.8 | 26 | 35.1 |  | 61 | 39.6 |
|  | T only | 9 | 11.2 | 5 | 6.8 |  | 14 | 9.1 |
| **Pre-biopsy MRI** | Yes | 80 | 98.8 | 74 | 100 |  | 154 | 99.3 |
|  | No | 1 | 1.2 | 0 | 0 |  | 1 | 0.7 |

Table S2: Patient and tumour characteristics within the intervention and SOC arm, and overall. SOC = standard of care TP = Transperineal TR = Transrectal T&S = Targeted and systematic T only = Targeted only S only = Systematic only

|  |  | Predict  N(%) | SOC  N(%) | Overall  N(%) |
| --- | --- | --- | --- | --- |
| **Work status** | Employed | 18(22.2) | 16(21.3) | 34(21.8) |
|  | Not in paid employment | 1(1.2) | 2(2.7) | 3(1.9) |
|  | Retired | 48(59.3) | 43(57.3) | 91(58.3) |
|  | Self-employed | 13(16.0) | 13(17.3) | 26(16.7) |
|  | Not recorded | 1(1.2) | 1(1.3) | 2(1.3) |
| **Highest Educational level** | School | 22(27.2) | 22(29.3) | 44(28.2) |
|  | College | 28(34.6) | 23(30.7) | 51(32.7) |
|  | University | 11(13.6) | 12(16.0) | 23(14.7) |
|  | Post-graduate | 11(13.6) | 12(16.0) | 23(14.7) |
|  | Not recorded | 9(11.1) | 6(8.0) | 15(9.6) |
| **Relationship status** | Living with partner | 67(82.7) | 58(77.3) | 125(80.1) |
|  | Single | 4(4.9) | 14(18.7) | 18(11.5) |
|  | Widower | 2(2.5) | 2(2.7) | 4(2.6) |
|  | Not recorded | 8(9.8) | 1(1.3) | 9(5.8) |
| **Family status** | 1 or more children | 63(77.8) | 61(81.3) | 124(79.5) |
|  | No children | 11(13.6) | 13(17.3) | 24(16.0) |
|  | Not recorded | 7(8.6) | 1(1.3) | 8(5.1) |

Table S3: Social characteristics of participants

| **DMPQ Statement** | Predict  N(%) | SOC  N(%) | Overall  N(%) |
| --- | --- | --- | --- |
| *I prefer to make treatment decisions on my own* | 2(2.5) | 3(4.0) | 5(3.2) |
| *I prefer to make treatment decisions after hearing my doctor’s opinion* | 32(39.5) | 27(36.0) | 59(37.8) |
| *I prefer to make treatment decisions with my doctor* | 42(51.9) | 44(58.7) | 86(55.1) |
| *I prefer my doctor to make treatment decisions after talking to me* | 4(4.9) | 1(1.3) | 5(3.2) |
| *I prefer my doctor to make treatment decisions on his/her own* | 0(0) | 0(0) | 0(0) |
| *Not recorded* | 1(1.2) | 0(0) | 1(0.6) |

Table S4: Responses to the decision-making preference questionnaire. Choose statement the participant most agrees with.

| **Measure** | **observations** | **Control (n=74)**  **Mean (95% CI)** | **PREDICT (n=78)**  **Mean (95% CI)** | **P value** |
| --- | --- | --- | --- | --- |
| STAI Y1 Score (***State*** of anxiety) | 155 | 37.60 (34.98-40.22) | 35.49 (32.98-37.99) | 0.124 |
|  |  |  | 10 |  |
| STAI Y2 score (**Trait** of anxiety) | 155 | 34.03 (31.75-36.30) | 33.05 (30.73-35.37) | 0.275 |

Table S5: Comparison of total state trait anxiety inventory (STAI) scores for the control group and PREDICT group. Accepted values for: low (20-37), medium (38-44) and high (45-80).

|  | **Did PREDICT make you more or less likely to want radical treatment?** | | | | |
| --- | --- | --- | --- | --- | --- |
| **Final Treatment** | Less likely | No change | More likely | Unsure | Total |
| AS/Conservative | 23 | 16 | 1 | 6 | 46 |
| Surgery | 3 | 12 | 8 | 2 | 25 |
| Radiotherapy | 1 | 2 | 3 | 2 | 8 |
| Other | 1 | 1 | 0 | 0 | 2 |
| Total | 28 (34.6%) | 31 (37.8%) | 12 (14.8%) | 10 (12.3%) |  |

Table 6: Cross tabulation of whether Predict Prostate led to patients being more or less likely to want radical treatment, and their final treatment decision.

**Table S6**: Cross tabulation of participants’ treatment preferences and whether PREDICT would make them more likely to want radical treatment. Apparent trends are highlighted.

| **Current treatment preference** | **Control** | **PREDICT** | **Total** |
| --- | --- | --- | --- |
| **AS/Conservative** | 40 (53%) | 46 (57%) | 86 (55%) |
| **Surgery** | 21 (28%) | 21 (25%) | 42 (27%) |
| **Radiotherapy** | 8 (11%) | 7 (9%) | 15 (10%) |
| **Other** | 2 (3%) | 2 (2%) | 4 (3%) |
| **No preference** | 4 (5%) | 5 (6%) | 9 (6%) |
|  | 75 | 81 | 156 |

**Table S7.** Reported treatment preference in each arm of the study

|  | Control (%) | PREDICT (%) | Total |
| --- | --- | --- | --- |
| **AS/Conservative** | 45 (60.0) | 46 (56.8) | 91 (58.3) |
| **Surgery** | 21 (28.0) | 25 (30.9) | 46 (29.5) |
| **Radiotherapy** | 8 (10.7) | 10 (12.3) | 18 (11.5) |
| **Other** | 1 (1.3) | 0 (0.0) | 1 (0.6) |
| **Total** | 75 | 81 | 156 |

**Table S8:** Final treatment decision

|  | Did you find PREDICT useful? |
| --- | --- |
| Yes | 73 |
| Unsure | 8 |
| No | 0 |

|  | Would you recommend PREDICT? |
| --- | --- |
| Yes | 73 |
| Unsure | 5 |
| No | 0 |

**Figures S1 and S2**. Responses to the questions shown among men in the PREDICT group.

|  | **Yes** | **No** | **Unsure** |
| --- | --- | --- | --- |
| *Did you find PREDICT useful?* | 73 | 0 | 8 |
| *Would you recommend PREDICT?* | 76 | 0 | 5 |

Table S9: AS per figures C1 and C2

*excellent & helpful*

*excellent information. Very helpful*

*good information given and will help me make the correct decision for me*

*good tool*

*great tool - great user interface - well designed and presented by Sam*

*helpful*

*Helpful in understanding the numbers of people affected. Presentation was excellent and informative.*

*I found the tool very informative and with good output options (visuals and text)*

*I have found the session / using the tool very helpful*

*I think these surveys are very important and should be done all the time*

*it helps to make a decision*

*it seems good to me*

*it was interesting and informative*

*It was very informative and gave me self assurance that I was making the correct decision on treatment*

*keep things as simple as possible*

*PREDICT my impression was it might be a tool to predict my only option for treatment - maybe "PREDICT" prostate is not the right wording?*

*Received a good explanation as to the aims & objectives of the programme*

*thank you*

*the dots image was very good for understanding the data*

*this is a very useful and enlightening tool*

*This tool will be helpful to others who have an enquisitive mind. It was helpful to see the diagrams.*

*very clear webstie/model. Very helpful*

*very good information provider*

*very helpful*

*very helpful*

*very helpful*

*very informative*

*very informative & has put me at ease*

*Very useful session which did help confirm and alleviate my concerns*

*very useful tool& questionnaire Question 2 would be more comprehensive if it recognises the role of spouse / partner in decision making*

**Table S10**: Free text responses of participants in the PREDICT arm. Response to the question ‘any other comments or feedback?’

Additional Files

Additional File 1: Prescribed terminology for researchers presenting Predict Prostate.

Additional File 2: Complete study questionnaire

Additional File 3: Study protocol
