## Additional File 1 - Study appointment protocol for "Clinical impact of the *Predict* Prostate risk communication tool in men newly diagnosed with non-metastatic prostate cancer: a multi-centre randomised controlled trial"

**PREDICT Prostate Patient Study – Study Appointment Protocol**

1. Reintroduce study generally
2. Go over PIS and answer any outstanding questions patient may have
3. Go over consent form and ensure it’s all signed appropriately
   (reiterate that only half will be randomised to see the Predict model)
4. Ask about previous access to the PREDICT Prostate website – and complete section in CRF accordingly.
5. Open brown envelope to check randomisation group
6. If randomised to questionnaire only: Provide the questionnaire, **crossing through or removing pages 9 and 10.** After completion, thank the patient for their time and inform them their participation in the study has completed.
7. If randomised to PREDICT Prostate + questionnaire follow the protocol/script below:
   a. Explain they have been randomised to see the model
   b. Open [prostate.predict.nhs.uk](https://wintoncentre.maths.cam.ac.uk/files/predict/prostate2/" \l "/)
   c. Press F11 to make the model full screen
   d. Start on the ‘Home’ page and read through the 3 paragraphs
   e. Move to the ‘About Predict Prostate’ page and skim/summarise the 3 paragraphs (emphasise that the model does not itself give advice, it simply provides some information, it is built around data from other men and is a ‘best guess’ of what outcomes might be)
   f. If the participant has more questions about the model itself – consider going through the ‘FAQ’ section.
   g. Go to the ‘Predict Prostate Tool’ page
   h. Enter the participants’ details
   i. Scroll to the ‘Results’ section
      i. Start on the ‘Icons’ section and select the ‘conservative’ management and ’15’ years tab
      ii. Explain that the icons represent 100 men with your characteristics. Explain that ‘Out of 100 men with the same age and disease characteristics as you, if all 100 had initial conservative management, the model estimates that XX would still be alive at 15 years, XX would have died from Prostate cancer and XX would have died from other causes. At this moment we cannot say to which group you will belong.”
      iii. Press the ‘Radical’ treatment regime tab
      iv. Explain that ‘Out of 100 men with the same characteristics as you, if all 100 had radical treatment, the model estimates that XX would still be alive at 15 years, XX would have died from prostate cancer and XX would have died from other causes. It estimates that XX extra men out of 100 might be alive at 15 years if all 100 were radically treated.’
      v. Switch to the ‘Charts’ section. Explain that the model estimates that at 10 years and 15 years, with initial conservative management (i.e. surveillance), XX% and XX% would be alive.
      vi. Explain that the dotted yellow line shows what proportion would be alive if the prostate cancer deaths were completely excluded (i.e if there was zero chance of dying from prostate cancer)
      vii. Press ‘Radical’ next to ‘Treatment Regime’
      viii. Explain that with radical treatment (i.e. Radical prostatectomy or radiotherapy) the model estimates that XX% and XX% would still be alive at 10 and 15 years.
      ix. Switch to the ‘Texts’, ‘Tables’ and ‘Curves’ results and explain that these are all showing the same estimates but in different ways.
      x. Answer any questions on these graphs or explain the results as necessary (may need to switch between 10 and 15 years to get the same outcomes)
      xi. **Do not offer advice or recommendations beyond explaining the model itself**. Explain that they can discuss things further with their consultant or specialist nurse later if necessary.
   j. Scroll down to the ‘Potential Harms of Treatment’ section
   k. Explain that alongside benefits from treatment, there is a risk of potential harms and these should also be considered in any decision.
   l. Explain the bullet points – i.e. that these estimates for harms are **not individualised** to you, and have been taken from studies in different centres.
   m. Explain that ‘If 100 men were all treated by conservative management/ prostatectomy/radiotherapy these studies suggest that **at 3 years**, XX% would have Erectile dysfunction/Incontinence/Bowel dysfunction (using the shown definitions).
   n. Signpost to other sources of information and the websites listed.
   o. Read out the ‘Important’ note at the side/bottom of the page.
8. Provide the questionnaire, (**including pages 9 and 10).** After completion, thank the patient for their time and inform them their participation in the study has completed.
9. Thank the patient for their participation.

**Further information and contact details**

Mr David Thurtle, Academic Urology Group, Box 279, Addenbrooke’s Hospital, Hills Road, Cambridge, CB2 0QQ.
