## Additional File 2 - Patient Questionnaire for "Clinical impact of the *Predict* Prostate risk communication tool in men newly diagnosed with non-metastatic prostate cancer: a multi-centre randomised controlled trial"

**[Local Site name/logo]**
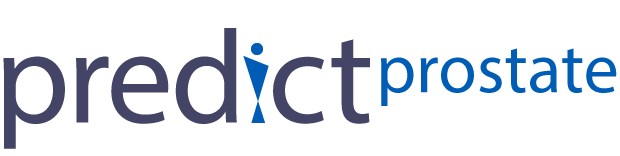

**PREDICT: *Prostate* Patient Study**

**Patient Questionnaire**

Thank you for agreeing to take part in this study.
Please complete this questionnaire to the best of your ability.

**Participant Details** (to be completed by Researcher)

Study ID _______________________________________

Questionnaire date______________________________

Study arm _____________________________________

1 A. Please answer the following questions about your own age and medical history

a) **Age**	__________________________(years)
b) **Other medical problems** _____________________________________________________________________________________________________________________________________
c) **Have you been admitted to hospital in the last 2 years?**	Yes	No
d) **If so, what was this for?** ______________________________________________
   __________________________________________________________________
   __________________________________________________________________

1 B. Please circle the appropriate answers below, or leave blank if you would prefer not to answer.

a) **Work status**	Retired	Employed	Self-employed	Not in paid employment
b) **Highest education level**	School	College	University	Post-graduate
c) **Relationship status**	Living with partner	Single	Widower
d) **Family status**	No children	1 or more children	Still planning children

2. Please choose which of the following statements you most agree with:

1. I prefer to make treatment decisions on my own.
2. I prefer to make treatment decisions after hearing my doctor's opinion.
3. I prefer to make treatment decisions together with my doctor
4. I prefer my doctor to make treatment decisions after talking to me.
5. I prefer my doctor to make treatment decisions on his/her own.

3. Please fill in the following questions about your current treatment preferences.

A. At this moment in time, do you have a preference to one of the following treatment options?

a) Surgery (prostatectomy)
b) Radiotherapy
c) Active Surveillance / Conservative Management
d) Other (please state)	_______________________________________________
e) No preference

B. On a scale from 0 to 100 how certain are you in your preference above?

0 10	20	30	40	50	60	70	80	90	100

4. Considering the option you prefer in question 3, please answer the following questions:

|  | Strongly Agree | Agree | Neither agree nor disagree | Disagree | Strongly Disagree |
| --- | --- | --- | --- | --- | --- |
| 1. I know which options are available to me | □ | □ | □ | □ | □ |
| 2. I know the benefits of each option | □ | □ | □ | □ | □ |
| 3. I know the risks and side effects of each option | □ | □ | □ | □ | □ |
| 4. I am clear about which benefits matter most to me | □ | □ | □ | □ | □ |
| 5. I am clear about which risks and side effects matter most to me | □ | □ | □ | □ | □ |
| 6. I am clear about which is more important to me (the benefits or the risks and side effects) | □ | □ | □ | □ | □ |
| 7. I have enough support from others to make a choice | □ | □ | □ | □ | □ |
| 8. I am choosing without pressure from others | □ | □ | □ | □ | □ |
| 9. I have enough advice to make a choice | □ | □ | □ | □ | □ |
| 10. I am clear about the best choice for me | □ | □ | □ | □ | □ |
| 11. I feel sure about what to choose | □ | □ | □ | □ | □ |
| 12. This decision is easy for me to make | □ | □ | □ | □ | □ |
| 13. I feel I have made an informed choice | □ | □ | □ | □ | □ |
| 14. My decision shows what is important to me | □ | □ | □ | □ | □ |
| 15. I expect to stick with my decision | □ | □ | □ | □ | □ |
| 16. I am satisfied with my decision | □ | □ | □ | □ | □ |

**4. For each of the following, please select how important they are to you when deciding about your treatment.**

a) **The chance of dying from prostate cancer**

Not important	Slightly important	Moderately important	Important	Very important

b) **The risk of urinary problems**

Not important	Slightly important	Moderately important	Important	Very important

c) **The risk of bowel problems**

Not important	Slightly important	Moderately important	Important	Very important

d) **The risk of sexual problems**

Not important	Slightly important	Moderately important	Important	Very important

e) **The burden of the treatment itself**

Not important	Slightly important	Moderately important	Important	Very important

f) **The thought of living with a cancer that is untreated**

Not important	Slightly important	Moderately important	Important	Very important

5. STAI Form Y-1

A number of statements which people have used to describe themselves are given below. Read each statement and then circle the appropriate value to the right of every statement to indicate how you feel **at this moment**. There are no right or wrong answers. Do not spend long on any one statement but give the answer which seems to describe your **present feelings** best.

|  | Not at all | Some-what | Moder-ately so | Very much so |
| --- | --- | --- | --- | --- |
| 1. I feel calm | 1 | 2. | 3. | 4. |
| 2. I feel secure | 1 | 2. | 3. | 4. |
| 3. I am tense | 1 | 2. | 3. | 4. |
| 4. I feel strained | 1 | 2. | 3. | 4. |
| 5. I feel at ease | 1 | 2. | 3. | 4. |
| 6. I feel upset | 1 | 2. | 3. | 4. |
| 7. I am presently worrying over possible misfortunes | 1 | 2. | 3. | 4. |
| 8. I feel satisfied | 1 | 2. | 3. | 4. |
| 9. I feel frightened | 1 | 2. | 3. | 4. |
| 10. I feel comfortable | 1 | 2. | 3. | 4. |
| 11. I feel self-confident | 1 | 2. | 3. | 4. |
| 12. I feel nervous | 1 | 2. | 3. | 4. |
| 13. I am jittery | 1 | 2. | 3. | 4. |
| 14. I feel indecisive | 1 | 2. | 3. | 4. |
| 15. I am relaxed | 1 | 2. | 3. | 4. |
| 16. I feel content | 1 | 2. | 3. | 4. |
| 17. I am worried | 1 | 2. | 3. | 4. |
| 18. I feel confused | 1 | 2. | 3. | 4. |
| 19. I feel steady | 1 | 2. | 3. | 4. |
| 20. I feel pleasant | 1 | 2. | 3. | 4. |

6. STAI Form Y-2
The next set of questions relate to **how you generally feel.** Read each statement and then circle the value to the right of every statement that indicates how you generally feel. There is no right or wrong answer. Do not spend too much time on any one statement but give the answer which seems to describe **how you** **generally feel**.

|  | Not at all | Some-what | Moder-ately so | Very much so |
| --- | --- | --- | --- | --- |
| 21. I feel pleasant | 1 | 2. | 3. | 4. |
| 22. I feel nervous and restless | 1 | 2. | 3. | 4. |
| 23. I feel satisfied with myself | 1 | 2. | 3. | 4. |
| 24. I wish I could be as happy as others seem to be | 1 | 2. | 3. | 4. |
| 25. I feel like a failure | 1 | 2. | 3. | 4. |
| 26. I feel rested | 1 | 2. | 3. | 4. |
| 27. I am calm, cool, and collected | 1 | 2. | 3. | 4. |
| 28. I feel that difficulties are piling up so that I cannot overcome them | 1 | 2. | 3. | 4. |
| 29. I worry too much over something that really doesn’t matter | 1 | 2. | 3. | 4. |
| 30. I am happy | 1 | 2. | 3. | 4. |
| 31. I have disturbing thoughts | 1 | 2. | 3. | 4. |
| 32. I lack self confidence | 1 | 2. | 3. | 4. |
| 33. I feel secure | 1 | 2. | 3. | 4. |
| 34. I make decisions easily | 1 | 2. | 3. | 4. |
| 35. I feel inadequate | 1 | 2. | 3. | 4. |
| 36. I am content | 1 | 2. | 3. | 4. |
| 37. Some unimportant thoughts run through my mind and bothers me | 1 | 2. | 3. | 4. |
| 38. I take disappointment so keenly that I can’t put them out of my mind | 1 | 2. | 3. | 4. |
| 39. I am a steady person | 1 | 2. | 3. | 4. |
| 40. I get in a state of tension or turmoil as I think over my recent concerns and interests | 1 | 2. | 3. | 4. |

7. The next 3 questions are very difficult questions to answer. Please make your best estimate or guess for each question below, selecting any value between 0 and 100.

a) Out of 100 men with the exact same characteristics as you, how many do you think would **die from prostate cancer** in 15 years if not treated?

0	10	20	30	40	50	60	70	80	90	100

b) Out of 100 men with the exact same characteristics as you, how many do you think would **die from other causes** (i.e. NOT prostate cancer) in 15 years?

0	10	20	30	40	50	60	70	80	90	100

c) Out of 100 men with the exact same characteristics as you, how many **extra** men do you think would be alive at 15 years if all 100 men were treated by surgery or radiotherapy?

0	10	20	30	40	50	60	70	80	90	100

**Additional Questions A1-A8 for men who have seen PREDICT: Prostate only**

Having seen the PREDICT: *Prostate* estimates, please select from the following statements:

A1. The number of men estimated to die **from prostate cancer** following conservative management were:

Less than I expected	Similar to what I expected	More than I expected

A2. The number of men estimated to die **from other causes** (i.e. not prostate cancer), following conservative management, were:

Less than I expected	Similar to what I expected	More than I expected

A3. The number of **extra** men alive following radical treatment compared to conservative management were:

Less than I expected	Similar to what I expected	More than I expected

A4. Did you find PREDICT: *Prostate* to be helpful?

Yes	No	Unsure

A5. Would you recommend using the tool to other men in your position?

Yes	No	Unsure

A6. Did PREDICT: *Prostate* make you feel more or less likely to want radical treatment (surgery or radiotherapy) for your prostate cancer?

More	likely	Less likely	No change	Unsure

A6. Is there anything about the model that you did not like, or would change?

__________________________________________________________________________
__________________________________________________________________________
__________________________________________________________________________

A7. Is there anything else you would like to see added to the PREDICT: Prostate model/website?
________________________________________________________________________
________________________________________________________________________________________________________________________________________________

A8. Any other comments or feedback?
________________________________________________________________________
________________________________________________________________________________________________________________________________________________
