## Additional File 3 - Study Protocol for "Clinical impact of the *Predict* Prostate risk communication tool in men newly diagnosed with non-metastatic prostate cancer: a multi-centre randomised controlled trial"

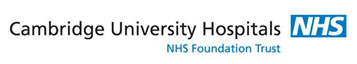

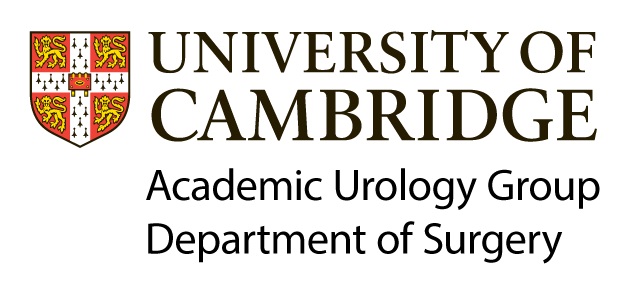

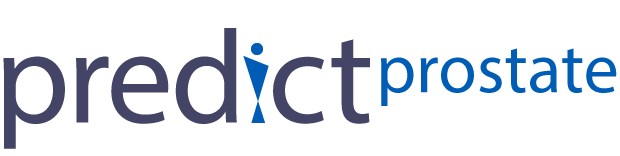

Academic Urology Group, Normal Bleehan Oncology Offices 1^st^ Floor, The Former Clinical Workshops, Box 193, Addenbrooke’s Hospital, Hills Road, Cambridge, CB2 0QQ

**Mr. David Thurtle** BMedSci BMBS MRCS AFHEA

*Clinical Research Associate*

*Honorary Urology Registrar*

**Mr. Vincent J. Gnanapragasam** PhD FRCS FRCSEd(Urol)

*Lecturer in Uro-Oncology*

*Honorary Consultant Urological Surgeon*

**FULL/LONG TITLE OF THE STUDY**

Evaluation of a new tool, PREDICT: *Prostate,* to aid treatment decision-making for men with newly diagnosed non-metastatic prostate cancer

**SHORT STUDY TITLE / ACRONYM**

PREDICT: *Prostate* Patient Study

**PROTOCOL VERSION NUMBER AND DATE**

1.3 March 2019

**RESEARCH REFERENCE NUMBERS**

| **IRAS Number:**  **REC Reference:** | 249699  **18/EE/0254** |
| --- | --- |
| **SPONSORS Number:** | Cambridge University Hospitals NHSFT: A094875  University of Cambridge #TBC |
| **FUNDERS Number:** | The Urology Foundation Research Scholarship  (RG #96622) |

### SIGNATURE PAGE

The undersigned confirm that the following protocol has been agreed and accepted and that the Chief Investigator agrees to conduct the study in compliance with the approved protocol and will adhere to the principles outlined in the Declaration of Helsinki, the Sponsor’s SOPs, and other regulatory requirement.

I agree to ensure that the confidential information contained in this document will not be used for any other purpose other than the evaluation or conduct of the investigation without the prior written consent of the Sponsor

I also confirm that I will make the findings of the study publically available through publication or other dissemination tools without any unnecessary delay and that an honest accurate and transparent account of the study will be given; and that any discrepancies from the study as planned in this protocol will be explained.

| **For and on behalf of the Study Sponsor:** | | |
| --- | --- | --- |
| Signature:  ...................................................................................................... |  | Date: ....../....../...... |
| Name (please print):  ...................................................................................................... |  |  |
| Position: ...................................................................................................... |  |  |
| **Chief Investigator:** | | |
| Signature:  ......................................... 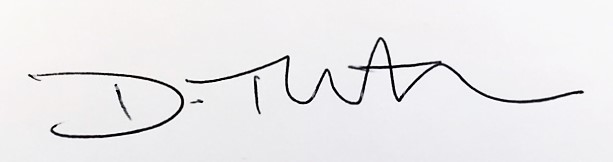……............ |  | Date:  ……/…../….. |
| Name:…………………..Mr David Thurtle……………....................  Position: Clinical Research Associate and Honorary Urology Registrar |  |  |

### LIST of CONTENTS

| **GENERAL INFORMATION** | **Page No.** |
| --- | --- |
| CONTACT DETAILS | 1 |
| TITLE PAGE | 2 |
| RESEARCH REFERENCE NUMBERS | 2 |
| SIGNATURE PAGE | 3 |
| LIST OF CONTENTS | 4 |
| KEY STUDY CONTACTS | 5 |
| STUDY SUMMARY | 5 |
| FUNDING | 7 |
| ROLE OF SPONSOR AND FUNDER | 7 |
| PROTOCOL CONTRIBUTORS | 7 |
| STUDY FLOW CHARTS | 8 |
| **SECTION** | |
| 1. BACKGROUND | 9 |
| 2. RATIONALE | 9 |
| 3. RESEARCH QUESTION/AIM(S) | 10 |
| 4. STUDY DESIGN/METHODS | 10 |
| 5. STUDY SETTING | 12 |
| 6. SAMPLE AND RECRUITMENT | 12 |
| 7. ETHICAL AND REGULATORY CONSIDERATIONS | 14 |
| 8. DISSEMINATION POLICY | 17 |
| 9. REFERENCES | 17 |
| 10. APPENDICES | 18 |

### KEY STUDY CONTACTS

| Chief Investigator | Mr David Thurtle, 07732097106, Ext 348441, |
| --- | --- |
| Study Co-ordinator | Mr David Thurtle 07732097106, Ext 348441, |
| Co-investigator / Supervisor | Mr Vincent Gnanapragasam 01223 348176,  |
| Study Co-Sponsors | Cambridge University Hospitals NHS Foundation Trust  Stephen Kelleher,  R&D Manager, Cambridge University Hospitals NHSFT, Hills Road, Cambridge, CB2 0QQ,  Tel: 01223 256 620, Fax: 01223 348494, E-mail:   University of Cambridge  Carolyn Read, Research Governance Officer, University of Cambridge School of Clinical Medicine, Addenbrooke’s Hospital, Hills Road, Cambridge CB2 0SP |
| Funder(s) | The Urology Foundation |
| Key Protocol Contributors | David Thurtle, 07732097106, Ext 348441,  Vincent Gnanapragasam, 01223 348176, |
| Committees | Cambridge Urology Translational Research and Clinical Trials (CU-TRACT), c/o Susan Platt, Team Administrator, 01223 348176 |

**STUDY SUMMARY**

| Study Title | Evaluation of a new tool, PREDICT: *Prostate,* to aid treatment decision-making for men with newly diagnosed non-metastatic prostate cancer |
| --- | --- |
| Internal ref. no. (or short title) | PREDICT: *Prostate* Patient Study |
| Study Design | Prospective randomised controlled study |
| Study Participants | Men diagnosed with non-metastatic prostate cancer suitable for active surveillance or radical treatment. |
| Planned Size of Sample (if applicable) | Target recruitment rate 150/year |
| Follow up duration (if applicable) | 1 year |
| Planned Study Period | August 2018 – July 2020 |
| Research Question/Aim(s) | Does PREDICT: *Prostate* improve treatment decisional confidence and anxiety amongst men diagnosed with non-metastatic prostate cancer?  Does PREDICT: *Prostate* affect patient decision-making with regards to treatment for localised prostate cancer?  How do PREDICT: *Prostate* outcomes compare to patients’ perceptions about long term survival outcomes. |
| Lay summary | The number of men diagnosed with prostate cancer is rising, in the vast majority of cases the disease has not spread elsewhere (non-metastatic). Here, treatment decisions are complex, with the risks of a cancer diagnosis balanced against potential problems associated with treatment. National guidelines advise that evidence-based decision aids should be used, yet no adequate individualised decision aid yet exists. To address the absence of such an important aid we have developed ‘PREDICT: *Prostate’.* This is a decision model and website which provides personalised survival estimates based on an individual’s characteristics and those of their cancer. The model allows the risk of dying from cancer to be contextualised against other risks of death and estimates the potential survival benefit from treatment.  This study seeks to assess the clinical usefulness and potential impact of PREDICT: *Prostate* amongst patients diagnosed with prostate cancer. We will assess the impact of the model on treatment decision-making, and on levels of concern, confidence and anxiety amongst newly diagnosed men. We will also assess how PREDICT estimates compare to patients’ perceptions about survival. We will also seek feedback about the model and its usefulness.  As per the study flow chart below; after selection, recruitment and informed consent, patients will be assigned to either the ‘standard of care’ (SOC) arm or the ‘SOC + PREDICT’ arm of the study. Prior to their next clinical follow-up appointment, participants will be invited to attend the hospital for a study meeting. During this meeting, all participants will be asked to complete a questionnaire. For those in the ‘SOC and PREDICT’ arm, this questionnaire will follow a semi-structured presentation of the PREDICT: *Prostate* model; those in the SOC arm will complete the questionnaire only. The participant’s involvement in the study will finish after completion of the questionnaire with no further intervention or involvement required. |

**FUNDING AND SUPPORT IN KIND**

| **FUNDER(S)**  (Names and contact details of ALL organisations providing funding and/or support in kind for this study) | **FINANCIAL AND NON FINANCIALSUPPORT GIVEN** |
| --- | --- |
| The Urology Foundation 1-2 St Andrews Hill London EC4V 5BY | Grant-funding for salary and study costs - through a peer-reviewed competitive national application process. |
| Any additional study costs will be internally funded, where necessary. |  |

**ROLE OF STUDY SPONSOR AND FUNDER**

The study will be sponsored by Cambridge University Hospitals NHS Foundation Trust and the University of Cambridge. The research and development (R&D) department of the sponsor will aide in study design and protocol-review.

The study will fall under the remit of the Cambridge Uro-Oncology Translational and Clinical Trials team (CU-TRACT) who will supervise the study’s conduct, progress and analysis. Final decisions regarding all aspects of the study will be taken by the CI in collaboration with the CU-TRACT team.

The project will be funded through a research grant received from The Urology Foundation. This funder will have no influence on the design, analysis or dissemination of the study. Any small additional costs relating to this study will be supported through departmental funding, via Mr Gnanapragasam

**PROTOCOL CONTRIBUTORS**

The protocol has been reviewed and critiqued by the sponsor following initial drafting by the study CI in collaboration with Mr Vincent Gnanapragasam, Academic Urology Group lead and CU-TRACT team leader. The protocol has been assessed and amended by members of the nursing and medical teams, and has been through an external peer-review process.

| **KEY WORDS:** | Prostate Cancer  Prognostic model  Impact study  Medical decision-making |
| --- | --- |

**PREDICT: Prostate Patient Study – Timeline.**

| Activity | 2018 | | | | 2019 | | | | 2020 | | | |
| --- | --- | --- | --- | --- | --- | --- | --- | --- | --- | --- | --- | --- |
|  | Q1 | Q2 | Q3 | Q4 | Q1 | Q2 | Q3 | Q4 | Q1 | Q2 | Q3 | Q4 |
| Study design and review |  |  |  |  |  |  |  |  |  |  |  |  |
| HRA Application and review |  |  |  |  |  |  |  |  |  |  |  |  |
| Patient recruitment |  |  |  |  |  |  |  |  |  |  |  |  |
| Data analysis |  |  |  |  |  |  |  |  |  |  |  |  |
| One-year review and write-up |  |  |  |  |  |  |  |  |  |  |  |  |
| Final write-up |  |  |  |  |  |  |  |  |  |  |  |  |
| External presentations/ publications |  |  |  |  |  |  |  |  |  |  |  |  |

**PREDICT: Prostate Patient Study Flow Chart**

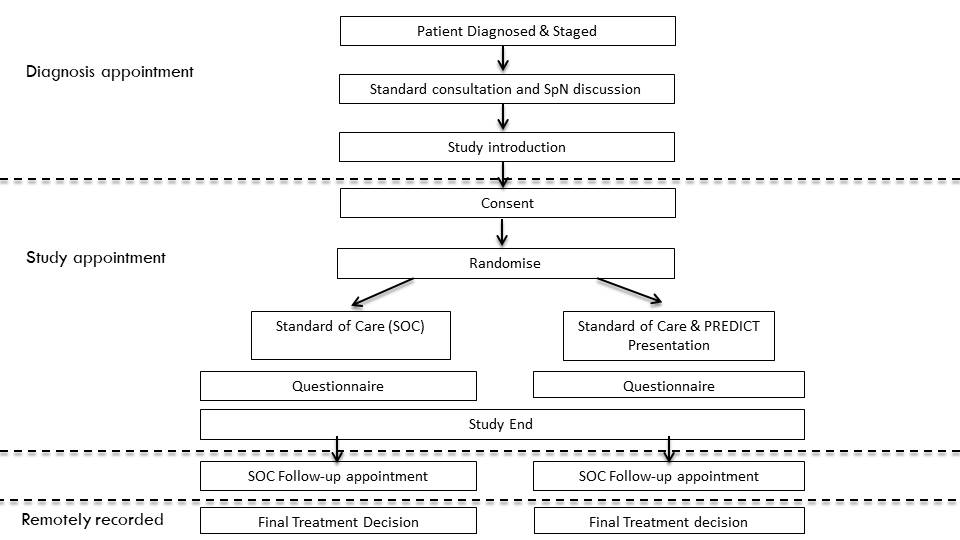

**STUDY PROTOCOL**

**TITLE**

Evaluation of a new tool, PREDICT: *Prostate,* to aid treatment decision-making for men with newly diagnosed non-metastatic prostate cancer

### 1 BACKGROUND

Prostate cancer (PCa) is the commonest cancer affecting males and is a leading cause of cancer-related morbidity [1]. The vast majority of new presentations (>80%) are with localised or locally advanced disease representing a significant healthcare and economic burden [2]. Treatment decisions are notoriously complex with the risk of progression and psychological impact of a cancer diagnosis balanced against potential morbidity associated with radical treatment for indolent tumours. Prognostic stratification is therefore the cornerstone of management. However, no high-quality individualised model for survival exists as demonstrated by the inability of the American Joint Committee on Cancer (AJCC) to endorse a single prognostic model for non-metastatic PCa [3].

We have therefore developed a novel individualised prognostic model called ‘PREDICT: *Prostate*’ for non-metastatic PCa. This contextualises the relative PCa-specific and overall survival outcomes for men with newly diagnosed disease and allows modelling of the estimated impact of radical treatment on these outcomes. To develop this we used data from 10,089 men diagnosed with PCa in Eastern England with median follow-up of 9.8 years and 3,829 deaths (1,202 PCa-specific). The model demonstrated good discrimination within a UK validation cohort with Concordance-index 0.83 (95%CI: 0.80-0.85) and 0.75 (95%CI: 0.74-0.77) for 15-year PCSM and Overall mortality respectively. This outperformed existing stratification criteria such as the European Association of Urology guidelines (C-index 0.69), National Cancer Collaborative Network criteria (C-index 0.72) and UCSF Cancer of the Prostate Risk Assessment (CAPRA) score (C-index 0.75). Accuracy has also been validated in external populations. The model has also been designed to meet all AJCC criteria for model adoption [4]. A publication on ‘PREDICT: Prostate’ is currently undergoing peer-review for publication.

### 2 RATIONALE

“An accurate prognostic model is of no benefit if it is not generalizable or doesn’t change behaviour” – Moons et al BMJ 2009 [5]

PREDICT: *Prostate* has many strengths in that it is built around real world data, the included predictors are unambiguously defined and the outcomes are transparent. Although accuracy appears favourable it is possible to have developed an accurate model that offers minimal clinical benefit. Although the model will be free to use, many stakeholders will expect to see evidence of clinical utility prior to adoption. In assessing the clinical utility we hope to quantify any potential benefit of using the model compared to current standard practice.

The underlying assumption that accurate outcome estimates lead to improved patient decision-making requires testing. ‘Impact studies’ such as this, seek to quantify whether using a prognostic model improves decision-making within a comparative design. Impact studies should by definition include a control group who receive standard care. “Only an impact analysis can determine whether use of the model is better than usual care” [5]. Alongside this, impact studies can be useful to study issues that may affect acceptability and uptake of a model in regular care – including usability. The ideal assessment outcome of survival is not a viable option, therefore we seek to assess the effect the model has on shorter term outcomes such as decision-certainty, decision-anxiety, decision-making behaviours and perceptions of disease severity.

### 3 RESEARCH AIMS

In this study we will seek to answer the following questions:

- Does PREDICT: *Prostate* improve decision confidence, and reduce anxiety, amongst men diagnosed with non-metastatic prostate cancer?
- Does PREDICT: *Prostate* change patient decision-making around treatment for non-metastatic prostate cancer?
- How do PREDICT: *Prostate* survival estimates compare to patients’ perceptions?

**3.1**	**Objectives**

1. To assess patient decisional certainty with and without use of the PREDICT: *Prostate* tool.

2. To gain insights into patient perceptions on PCa risks and the factors behind treatment decision-making, and assess whether certain patients may benefit more from using PREDICT: *Prostate.*

**3.2	Outcome measures**

**Primary outcome measure**

1. Patient scores on decisional certainty measured by the decisional conflict scale (DCS) [6]

**Secondary outcome measures**

1. Patient scores on state-anxiety measured on the State-Trait Anxiety Inventory (STAI-Y) [7]

2. Reported treatment preference and confidence in their decision on a 0-100 scale.

3. Actual treatment decided upon or received (as recorded in the medical notes).

### 4 STUDY DESIGN and METHODS of DATA COLLECTION AND DATA ANALYIS

The study is a prospective randomised study without medical or surgical intervention. The study will be introduced by a member of the clinical team following the diagnosis of non-metastatic prostate cancer. Prior to their next hospital appointment as part of their routine clinical pathway, participants will be invited to attend a study appointment at the hospital. After further discussion and informed consent, patients will be randomised to the standard of care (SOC) arm or the ‘SOC and PREDICT’ (intervention) arm of the study. During this study appointment, participants will be asked to complete a questionnaire. For those in the intervention arm, this questionnaire will follow a semi-structured presentation of the PREDICT: *Prostate* model. Patients on the SOC arm will simply be asked to complete the questionnaire. These questionnaires will assess treatment preferences, decisional confidence and anxiety, and patients’ perceptions of risk regarding their disease – using validated scores for each.

The PREDICT: *Prostate* model itself will be presented on a computer by an individual researcher or designated individual trained in the background of the model. They will present the model in a semi-structured manner. First the rationale, goals, and a detailed description of the decision aid will be presented, following the web-pages of the model. The patient’s individual details will be entered into the model and the results explained to them using positive and negative terms, and expressing uncertainty. For example “out of 100 patients with the same age and disease characteristics as you, 16 are expected to die from prostate cancer in the next 10 years, 10 are expected to die from other causes, and 74 are expected to still be alive. At this moment we cannot say to which group you will belong.” Graphs, charts, text, icons and actual numbers will be presented to the participants showing the estimated outcomes with conservative management and radical treatment – following the design of the website. Adverse effects information will also be presented through the website, alongside an explanation of their providence. The researcher will not themselves go into further details on technical aspects and will not offer clinical advice beyond explaining the website. All patients will be seeing a clinician in a follow-up appointment as part of their clinical pathway soon after completing the questionnaire, and have access to a specialist nurse at any point. If patients have new questions as a result of the questionnaire or from seeing PREDICT: Prostate they will have the opportunity to discuss these with their health care professionals through these mechanisms.

The webtool can be accessed for review using the following address:

[www.prostate.predict.nhs.uk](http://www.prostate.predict.nhs.uk)

**Data collection**

Case report forms (CRF) will be completed by a researcher for all study participants at study entry. Questionnaires will be completed by all study participants on the day of the first follow-up appointment. Copies of these questionnaires, which have been reviewed and edited by a patient and public involvement group are included in the appendix.

Impact of the decision aid may depend upon patient and tumour characteristics that affect their understanding, and perceptions about disease management. Therefore, data will be collected on patient demographic and tumour details. Details of tumour characteristics will be entered using details from the hospital electronic record system. The importance of various factors in decision-making will also be assessed such as survival, bowel function, urinary function and burden of treatment itself. Methodology for this purpose has been published previously [8].The scoring systems used within the questionnaire are validated scales that have been used widely in clinical research, namely the Decisional Conflict Scale (DCS) [6], Decision Making Preference Questionnaire (DMPQ) [9] and the State-Trait Anxiety Inventory (STAI-Y) [7]. The final treatment decision will be recorded from the medical notes.

**Data analysis**
We will use descriptive statistics to describe the participant cohort and check for imbalance between arms of the study. The impact of the intervention will be tested in an intention-to-treat analysis comparing the intervention group to the control group. Mean DCS and STAI scores between groups will be compared using independent samples t-test. Data analysis will be performed using Stata™ 14. Calculations for sample size are shown below. Subgroup analyses will be performed using responses to the DMPQQ, the patient’s prostate cancer risk group, and according to responses about the importance of associated issues in a patient’s decision-making. If further sites are added in future amendments, sub-group analysis will be performed by site to assess whether the location of the study had any impact on outcomes and to assess the variance**.** Qualitative feedback, patient comments and answers to questions about usefulness of the model will be collated and analysed using a deductive approach using our questions as a guide for grouping and analysing our data.

### 5 STUDY SETTING

This study will be implemented in prostate diagnostic clinics. The lead site will be Cambridge University Hospitals NHS Foundation Trust (CUH). Cambridge itself has strong research infrastructure, with the potential for collaborations across specialties. Patients at CUH are used to being invited into research projects, and are often keen to be involved in such studies. Testing this particular model within the East of England is particularly appropriate, given the model was developed using data from men diagnosed with PCa in the East of England.

This study falls perfectly within the construct of our current clinical pathway, without any disruption to patients or services. The study supplements the current clinical pathway by seeking to better-inform patients, yet does not require any additional practical interventions. After being told of their PCa diagnosis, men are always counselled by the informing clinician and a specialist nurse. Men are then given written information and links to useful websites to take home and think over. Men are then invited to meet with an oncologist and or a surgeon to discuss radiotherapy and surgery respectively before usually meeting with the diagnosing urologist again. These follow-up appointments provide opportunities for the questionnaire and intervention to be performed within the standard care pathway without expecting patients to make additional trips into hospital. Otherwise, if patients are willing, study appointments can be arranged at other mutually convenient times prior to their next follow-up appointment.

**6	SAMPLE AND RECRUITMENT**

**6.1	Eligibility Criteria**

**6.1.1	Inclusion criteria**

- Men newly diagnosed with primary non-metastatic PCa.
- Men for whom either active surveillance or radical treatment (prostatectomy +/- radiotherapy) are felt to be appropriate by the diagnosing clinician.
- Age 35-80 years
- Able to understand and sign the written Informed Consent Form

**6.1.2	Exclusion criteria**

- Subject is known to have a condition, which affects their ability to see, read or understand the decision aid
- Subject is known to have any other condition, which in the opinion of the investigator makes the subject unsuitable for study participation.
- The subject is unable to comprehend English. (PREDICT Prostate is only available in English currently)

**6.2	Sampling**

**6.2.1	Size of sample**

This study is both a qualitative and quantitative study aiming to explore the impact of PREDICT: Prostate in the decision-making pathway and patients’ impressions on the model. Sample size calculations are directed by the hypothesis that PREDICT: Prostate will improve decision-certainty, and reduce decisional conflict. Comparison between the intervention and control group will be performed by comparison of means. Decision certainty will be derived on a 0-100 scale from the DCS and the uncertainty sub-scale of the DCS. Using a conservative estimate of 20% change in decision certainty, (with SD 20, α 0.05, β 0.80) the minimum required sample size is 16 for each arm (i.e 32 total patients). However, some men may feel unable to state a treatment preference or complete the relevant parts of the questionnaire.

This effect size is an estimate. Similar studies using decision interventions have reported changes in actual treatment choices in excess of 20% [10], whilst others have reported effect sizes up to 38% for decision regret in a similar randomised controlled trial of a decision aid [11]. Therefore an effect size of 20% on reported decision-certainty appears reasonable.

It is estimated about 200 patients are diagnosed with non-metastatic PCa at Addenbrooke’s hospital each year. Therefore our recruitment target of 50 per year is a realistic one, and will be sufficiently powered to answer our primary question.

6.2.2 **Sampling technique / Randomisation**

Participants will be randomised to standard of care or standard of care & presentation of the PREDICT: Prostate model. Consecutive eligible patients will be approached wherever possible. Randomisation will be achieved by block random allocation (with random block sizes from 4-6) to achieve a greater equivalence between treatment groups in group sizes and baseline characteristics [12].

**6.3 Recruitment**

**6.3.1 Recruitment**

Eligible patients newly diagnosed with PCa will be informed about the study by their diagnosing clinician, specialist nurse or another member of their clinical care team. If interested in the study potential participants will then be approached in clinic and given information sheets, verbal information about the study and a copy of the consent form. Each will be given the opportunity to ask questions. Verbal consent to be contacted by telephone will be sought. Potential participants will then be contacted and invited to attend a study appointment at some point prior to their next clinical follow-up appointment – where formal written consent will be completed prior to the presentation of PREDICT or completion of questionnaires.

**6.3.2 Sample identification**

Eligible subjects will be identified from prostate diagnostic clinics and multi-disciplinary team meetings (MDTs). These patients will be identified by the PI, the patient’s responsible urology consultant or another member of the patient’s existing clinical care team. There will be no posters, adverts, websites or other active recruitment techniques. Patients will not be recruited through Patient Identification Centres.

No payments will be made to participants of the study. Participation will not routinely require any additional travel, visits or diversion from the clinical pathway.

**6.3.3 Consent**

Patients unable to give informed consent, unable to read English or those who lack capacity are not eligible for the study. No efforts will be made to recruit these men.

Suitable subjects diagnosed through urology clinics will be approached and invited to participate in the study, or for permission to contact them with regards to the study, by their consultant or specialist nurse, or another member of the direct care team. The study will not be introduced until after the diagnosis of non-metastatic prostate cancer has been given to them by the clinician. Men will be offered a comprehensive information sheet and a copy of the consent form. A discussion will be had about the nature and objectives of the study and possible risks associated with participation, between the potential participant and the CI, PI or an appropriately trained individual knowledgeable about the research named on the delegation log. Potential participants will be given the opportunity to raise any questions.

Consent will be taken at the start of the study appointment – allowing at least a 24 hour cooling off period from when the study was first introduced. Consent would be taken to complete a questionnaire with or without a preceding semi-structured presentation of the PREDICT: *Prostate* tool.

If a participant, who has given informed consent, loses capacity to consent during the study, or decides to withdraw consent, the participant and all identifiable data collected would be withdrawn from the study.

### 7 ETHICAL AND REGULATORY CONSIDERATIONS

The predominant ethical aspect of this study is that the prognostic model may affect actual treatment decision-making, indeed this is a secondary outcome. However, this is a prognostic model/decision aid that has been developed according to established methodology which meets international adoption standards [4, 13] . It has been developed by an expert team in Cambridge, including Professor Paul Pharaoh (Cancer Epidemiology) and Mr Vincent Gnanapragasam (Academic Urology). It has been developed and validated within a primary cohort of men from Eastern England and has also been externally validated in a cohort of Singaporean men. Therefore, the generalisability to other men in the East of England is particularly valid.
Any informed decision-making is an improvement on current practice where decision aids are very rarely used – despite NICE guidance recommending their regular use [14]. Individualised tools providing long term survival estimates like the PREDICT: *Prostate* model are almost non-existent in current PCa patient-counselling. Therefore, this represents a probable improvement on the current decision-making process. Further to this, all patients will still have the standard of care consultations with clinicians, with this study having no impact on these meetings or consultations. Participants will each be seen by a specialist clinician after the tool is presented, with no final decisions being made after presentation of the model and all patients will have the opportunity to discuss the results of the tool with a specialised clinician in these routine clinical appointments.

**7.1	Research Ethics Committee (REC) and other Regulatory review & reports**

Before the start of the study, a favourable opinion will be sought from the local Research Ethics Committee and NHS REC for the study protocol, informed consent forms and patient information sheets. Specifically, we state that:

- Substantial amendments that require review by NHS REC will not be implemented until that review is in place and other mechanisms are in place to implement at site.
- All correspondence with the REC will be retained.
- It is the Chief Investigator’s responsibility to produce the annual reports as required.
- The Chief Investigator will notify the REC of the end of the study.
- An annual progress report (APR) will be submitted to the REC within 30 days of the anniversary date on which the favourable opinion was given, and annually until the study is declared ended.
- If the study is ended prematurely, the Chief Investigator will notify the REC, including the reasons for the premature termination.
- Within one year after the end of the study, the Chief Investigator will submit a final report with the results, including any publications/abstracts, to the REC.

**Regulatory Review & Compliance**

If this study is performed at other sites within the East of England, regulatory review across sites will be overseen by the CI.

**Amendments**

Should amendments need to be made to the REC application, the CI will contact the sponsor who will assess whether an amendment is substantial or non-substantial. The sponsor will submit a valid notice of amendment to the REC for consideration. Substantive changes will be clearly communicated by email to the sponsor’s R&D Department and the REC. Amendment history will be tracked by renumbering of the protocol for each set of amendments made. This numbering will commence from 0.1 for the first draft protocol, and 1.0 for the first approved protocol.

**7.2	Peer review**

All patient questionnaires, the PIS and ICF have been reviewed and corrected by a patient and public involvement panel. The protocol has undergone external peer-review by Ms C Etheridge (Lead Macmillan Urology Clinical Nurse Sprecialist, Ipswich Hospital) and Mr M Sut (Consultant Urological Surgeon, Peterborough Hospital following the standards outlined below from NIHR/CRN:

Peer review must be independent, expert, and proportionate:

a) **Independent**: At least two individual experts should have reviewed the study. The definition of independent used here is that the reviewers must be external to the investigators’ host institution and not involved in the study in any way. Reviewers do not need to be anonymous.
b) **Expert**: Reviewers should have knowledge of the relevant discipline to consider the clinical and/or service based aspects of the protocol, and/or have the expertise to assess the methodological qualitative aspects of the study.
c) **Proportionate**: Peer review should be commensurate with the size and complexity of the study. Large multicentre studies should have higher level (more reviewers with broader expertise and often independent review committee or board), and potentially international peer review.

A copy of the completed peer-review forms will be submitted alongside other documentation.

**7.3	Protocol compliance**

Accidental protocol deviations will be adequately documented on the relevant forms and reported to the CI and Sponsor immediately. Deviations from the protocol which are found to frequently recur will mandate immediate action.

**7.4	Data protection and patient confidentiality**

All investigators and study site staff will comply with the requirements of the General Data Protection Regulations 2018 with regards to the collection, storage, processing and disclosure of personal information and will uphold the Act’s core principles.

Specifically, patient demographic and personal information will be kept securely on Trust computers. Every subject enrolled into the study will be given a unique study identifier which will be used to refer to the patients during analysis. Hence all data will be linked-anonymised at source. Non-identifiable study data will be stored on a password protected encrypted hard-drive for the use of this study and kept in locked offices. Paper consent forms will be regularly archived and stored in the CI office. Access to data will only be with the permission of the study CI. The number of individuals with access to these data will be limited as much as possible to enable quality control, audit, and analysis.

The Investigators will make all study documentation and related records available should a competent authority inspection occur, or an IRB / IEC request a review. Personalised data will be destroyed within 12 months of the end of the study. Access to the research data generated by the study will be determined by the CI responsible for the study, who will act as the data custodian. Non-identifiable research data will be retained for a maximum of 5 years following study closure.

Wording related to the gathering, retention, and use of data on the patient information sheet has been informed by the ‘transparency wording for public sector sponsors’ guidance on the HRA website.

7.5	Indemnity

NHS indemnity will apply in the management and conduct of the research. Specifically this will cover any legal liability of the sponsor for harm to participants arising from the management of this research.
Any potential legal liability of investigators/collaborators arising from harm to participants in the conduct of the research will be covered by the Site. All individuals working on the project will also have independent professional indemnity cover.

As the primary employer of the CI, the University of Cambridge will provide insurance for the design of the study. Unique reference for this study is HVS/2017/2282. The University Insurance Office has advised that insurance for negligent and non-negligent harm to research subjects can be arranged if this study is approved by the NHS Ethics committee. Cover is provided under the University's Clinical Trials and/or Human Volunteer Studies policy, the insurers are Newline, the insurance policy reference is *B0823Q31000177/WD1600523* and the Limit of Indemnity under the policy is £10,000,000 for each and every claim.

No specific arrangements are necessary for payment of compensation in the event of harm to research participants as a result of this study.

**7.6	Access to the final study dataset**

The CI and co-investigators working on the study will have access to the final anonymised dataset. Co-investigators or collaborators may be provided with fully-anonymised data when necessary, but will never be provided with any identifiable or potentially identifiable data.

##### 8 DISSEMINATION POLICY

##### 8.1 Dissemination policy

Analysis and reporting of the results from this study will commence whilst the study is ongoing, and will continue after the study has finished. A final study report will be prepared following completion of the study, with a further report made available after analysis of all the data. The full study report will be made available for public access.
The data itself will remain under the ownership of the sponsor, and controlled by the CI. Publications relating to data gained from this project will be presented and published wherever possible. All such presentations and publications should name the CI, and sponsoring institutions. There are no plans to actively contact participants to inform them of the study’s output, however, future publications will be publicised through the University Department of Surgery website or the Cambridge Urology website.

**8.2	Authorship eligibility guidelines and any intended use of professional writers**

On the final study report and in any future publications authorship will be according to the criteria for individually named authors set forward by The International Committee of Medical Journal Editors. The CI will be named on any such publication, and the sponsors fully acknowledged.

14.	NICE, National Institute for Health and Care Excellence NICE Guidelines [CG175] Prostate cancer: diagnosis and treatment. January 2014.

##### 10. APPENDICES

**10.1	Appendix 1- Required documentation**

Patient Information Sheet (PIS)

Informed Consent Form (ICF)

Patient questionnaires

Case Report Forms (CRF)

Research CVs of all research team members

Peer review forms

Certificate of insurance (University)

**10.2**	**Appendix 2 – Schedule of Procedures**

**10.3**	**Appendix 3 – Amendment History**

| **Amendment No.** | **Protocol version no.** | **Date issued** | **Author(s) of changes** | **Details of changes made** |
| --- | --- | --- | --- | --- |
| 1 | 1.1 | Application: December 2018 | David Thurtle | Amendment to the study pathway to allow study appointments to take place at any point (after at least 24 hours cooling off period) prior to the subjects next clinical appointment. This is instead of the study appointment having to take place immediately before the routine follow-up appointment which was problematic for two main reasons: Firstly, that some clinic appointments are very early such as 8.30am such that to have a study appointment prior to this is not feasible. Secondly, that some subjects aren’t to be seen again in clinic for some time after the study is introduced.  Small changes have been made in the protocol to reflect this alteration. Namely in: the lay summary (p6), study design (p10-11) and study setting (p12) – see tracked changes version.  Minor wording changes have also been made to the PIS and ICF and renumbering throughout to version 1.1. |
| 2 | 1.2 | Feb 2019 | David Thurtle | Sample size amended to 150/year rather than 50/year. |
| 3 | 1.3 | March 2019 | David Thurtle | Predict Prostate logo added on page1.  URL updated on page 11. |
